# Beyond the Wrist: Finger-Worn Accelerometers Enhance Assessment of Post-Stroke Motor Performance

**DOI:** 10.1101/2025.10.22.25338411

**Authors:** Yunda Liu, Gloria Vergara-Diaz, Benito Lorenzo Pugliese, Randie Black-Schaffer, Grace Kim, Paolo Bonato, Sunghoon Ivan Lee

## Abstract

**Background:** Accurate and objective assessment of motor performance is critical for effective stroke rehabilitation. While wrist-worn accelerometers are widely accepted as a valid tool for evaluating upper-limb motor performance, they primarily capture arm and forearm movements, overlooking hand and finger activity. This limitation reduces their ability to detect changes in distal function, hindering the broader integration of wearable-based motor performance metrics into clinical practice.

**Objective:** To determine whether finger-worn accelerometers, which capture both proximal and distal movements of the upper limbs, offer a more comprehensive assessment of motor performance by comparing their convergent validity with that of wrist-worn accelerometers.

**Methods:** Bilateral accelerometer data were collected from 24 stroke survivors using finger-worn and wrist-worn devices as they performed unscripted daily activities in a simulated home environment. Motor performance metrics from both sensor locations were analyzed for correlations with the Fugl-Meyer Assessment for Upper Extremity (FMA-UE) and sensitivity to differences in motor performance across impairment levels.

**Results:** Finger-worn accelerometer metrics showed stronger correlations with FMA-UE scores than those from wrist-worn sensors, largely due to their ability to capture fine hand movements. Additionally, finger-worn sensors demonstrated greater sensitivity in detecting performance differences between mildly and moderately impaired individuals.

**Conclusions:** By capturing both proximal and distal movements, finger-worn accelerometers demonstrate stronger convergent validity with standardized measures of post-stroke motor impairment compared to wrist-worn accelerometers. These findings highlight their potential for providing a more comprehensive assessment of motor performance in stroke survivors.

## Introduction

Stroke is a leading cause of brain injury in the United States, affecting approximately 800,000 individuals annually, with 9.4 million reporting a history of stroke (1). Approximately 80% of survivors experience upper-limb motor impairments, often more severe in the limb contralateral to the brain lesion (2). These impairments reduce muscle function, limiting the ability to perform daily activities (3), which affects patients’ functional independence and quality of life (4, 5). Typically in the United States, stroke survivors undergo two to three weeks of intensive inpatient rehabilitation post-stroke (6), followed by less intensive outpatient therapy for variable lengths of time, which often extends into the chronic phase to maximize functional recovery and independence (3, 7).

Effective stroke rehabilitation relies on tailoring therapeutic strategies to each patient’s motor and functional status (8, 9). Rehabilitation specialists and researchers typically assess motor capacity—what patients are capable of doing— through clinical observations of patients’ behaviors in structured settings using tools like the Fugl-Meyer Assessment for Upper Extremity (FMA-UE) (10) and Action Research Arm Test (ARAT) (11). On the other hand, patients pursue rehabilitation to improve motor performance, the actual execution of daily activities outside the clinic (12). Over the past decade, wrist-worn accelerometers have emerged as a validated, objective tool for assessing real-world motor performance by capturing movement data from everyday life, offering a promising outcome measure to inform therapeutic decision-making (13).

Despite their promise, wrist-worn accelerometers have limitations that restrict their broader clinical adoption (14). These devices primarily capture movements of the proximal limb segments, such as the arm and forearm, while largely missing distal movements involving the wrist and hand (15, 16)—regions that are critical for performing activities of daily living (ADLs) (17). As a result, wrist-worn metrics may underestimate actual upper-limb use (18), limiting their ability to detect meaningful changes in distal motor function. This is especially problematic when rehabilitation efforts target improvements in fine motor skills, where wrist-worn accelerometers may lack the sensitivity needed to capture subtle yet clinically important changes. Moreover, they provide only a partial understanding of how motor capacity, particularly distal function, translates into patients’ motor performance environments.

To overcome the limitations of wrist-worn accelerometers, researchers have explored sensing technologies that capture hand and finger movements for a more comprehensive assessment of motor performance. For instance, Friedman et al. have developed a wearable system integrating a wrist-worn device embedding magnetometers and a magnetic ring on the index finger to estimate wrist and finger joint angles (19, 20). However, this system requires two components—the wrist sensor and finger ring—increasing the wearable burden for patients and is limited to capturing only hand and finger movements, omitting arm and upper-arm activity. Additionally, it is susceptible to environmental magnetic interference. A more promising approach uses a miniaturized accelerometer worn on the index finger (16, 21, 22). This standalone device simplifies use, captures both fine hand and gross arm movements, and enhances translational potential by generating motor performance metrics consistent with those validated for wrist-worn accelerometers. However, its validation has primarily been limited to healthy individuals and a small cohort of stroke survivors performing scripted ADLs in controlled laboratory settings.

In this study, we extend prior research by evaluating whether finger-worn accelerometers provide a more comprehensive evaluation of motor performance than wrist-worn accelerometers. Given that finger-worn accelerometers capture both proximal and distal upper-limb movements, while wrist-worn sensors primarily detect proximal movements, we hypothesize that finger-worn accelerometers offer a more accurate and holistic assessment. To test the hypothesis, we examine the convergent validity of finger-worn accelerometers against the FMA-UE, a validated measure of motor capacity encompassing both proximal and distal components, and compare it to that of wrist-worn accelerometers. Additionally, we assess the sensitivity of finger-versus wrist-worn accelerometers in detecting motor performance variations across participants with differing levels of impairment.

### Method

### Study Participants

Participants were recruited from Spaulding Rehabilitation Hospital at Harvard Medical School in Boston, USA. Recruitment was advertised through multiple channels, including flyers and advertisements in clinical areas, referrals from physicians and clinicians, and outreach via patient registries and related platforms. To be eligible for the study, participants must meet the following criteria: 1) be between 18 and 80 years old, 2) have experienced a stroke at least six months prior to the data collection, and 3) mild and partly moderate impairment in the affected upper-limb, indicated by an FMA-UE score greater than 35. This threshold for FMA-UE was chosen to ensure participants have sufficient finger motor ability for meaningful assessment with finger-worn sensors (23). The study was approved by the Institutional Review Board of Mass General and Brigham Hospital (IRB #2019P002612).

### Experimental Protocol

Subsequently, they were equipped with custom-designed three-axis accelerometers on their index fingers and commercially available three-axis accelerometers (Shimmer3, Shimmer Research Inc., USA) on their wrists, as illustrated in Figure 1. Both devices sampled acceleration data at 50 Hz. Participants were instructed to perform up to 11 ADLs, as detailed in Table 1, within a simulated apartment setting. These ADLs were chosen to cover a wide range of upper-limb movements relevant to daily life, including both gross arm and fine hand movements, consistent with prior studies focusing on validating wearable devices for upper-limb motor performance assessment (17, 23). Participants were not required to perform ADLs they did not routinely undertake in their daily lives. They received guidance on locating materials needed for the ADLs in the simulated environment (e.g., a grocery bag in the kitchen or a toothbrush in the bathroom) but performed the tasks independently without specific instructions from research staff to ensure naturalistic execution. Participants could perform each ADL in a standing or sitting position based on their motor abilities and preferences. The dataset also included natural transitional movements between ADLs, such as sit-to-stand transitions and brief walking episodes between locations.

**Figure 1.**
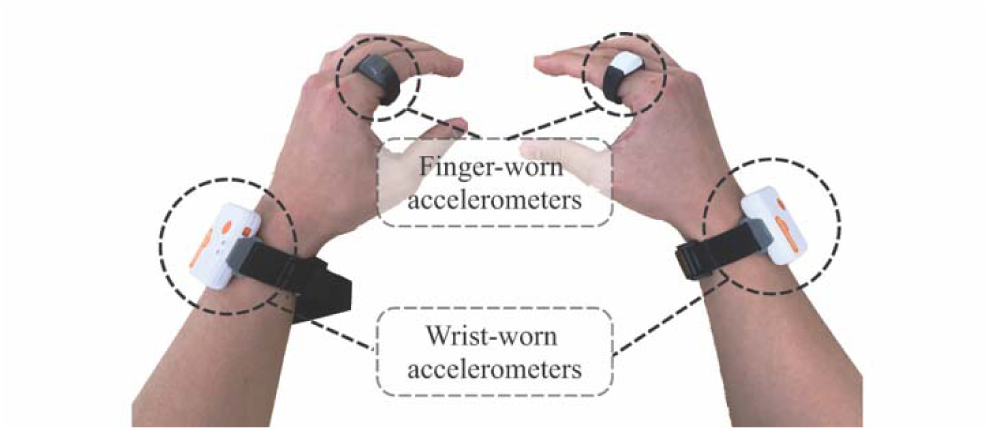
The finger-worn and wrist-worn accelerometers used in the data collection. During their clinical visit, participants underwent clinical assessment of their stroke-affected upper limb using the FMA-UE, which evaluates motor impairment on a 0 to 66 scale.

**Table 1.** Motor tasks resembling ADLs performed by study participants.

| # | Motor Task |
| --- | --- |
| 1 | Unloading a grocery bag |
| 2 | Placing a tablecloth on a table |
| 3 | Preparing a sandwich |
| 4 | Drinking water from a cup |
| 5 | Transferring the plate from the kitchen counter to a washing machine |
| 6 | Mopping the floor |
| 7 | Cleaning the table |
| 8 | Folding bath towels |
| 9 | Brushing teeth |
| 10 | Writing on paper |
| 11 | Donning/doffing a coat |

### Assessment of Motor Performance Using Wrist-Worn and Finger-Worn Accelerometers

We computed two well-established and validated measures of motor performance using three-axis acceleration data: intensity ratio and use ratio (22, 24). These metrics quantified differences in activity levels between the stroke-affected and contralateral limbs. Specifically, the intensity ratio evaluated the relative intensity of movement between the limbs, while the use ratio measured the comparative duration of active limb use.

To calculate these measures, the magnitude of the raw three-axis acceleration time-series from each sensor was computed: 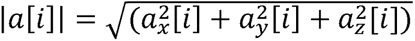, where *i* represents the index for each sample. To attenuate high-frequency, non-human-generated noise and remove gravitational component, the raw acceleration magnitude was band-pass filtered using a 4^th^ order Butterworth filter with cut-off frequencies of 0.1 Hz and 2 Hz (25, 26). The filtered acceleration magnitude time-series were then downsampled by averaging the data over one-second, nonoverlapping windows. The downsampled time-series for the stroke-affected and contralateral limbs are denoted as |*a_s_* [*t*]| and |*a_c_*[*t*]|, respectively, where *t* represents time in seconds. One-second window was chosen based on prior work to capture brief, intermittent upper-limb movement characteristics of ADLs in stroke survivors (22).

The intensity ratio *r_i_* was calculated by taking the log ratio between the acceleration magnitudes of the stroke-affected and contralateral limbs for each one-second window. The average value of this log ratio was then computed across the entire monitoring period:

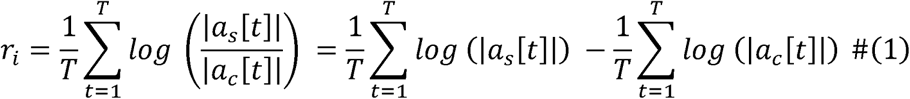

where *T* represents the total duration of the recorded data in seconds.

The use ratio *r_u_* was computed by taking the log ratio of the active use duration between the stroke-affected and contralateral limbs:

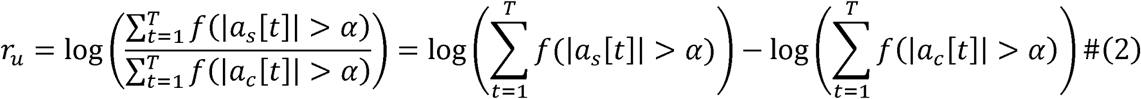

where *f*(.) is a boolean function that returns 1 if the input condition is true and 0 otherwise, and *α* is the minimal acceleration threshold for identifying active use within each epoch. While prior studies typically used a fixed of 3.33 × 10^−3^ *g* for wrist-worn accelerometers (23, 27), we treated as a tunable hyperparameter to determine active movements in a data-driven manner. Finger-worn sensors, due to the pendulum effect and their sensitivity to find hand movement, typically produced larger acceleration magnitudes than wrist-worn sensors. To account for these differences in movement profiles, *α* was independently optimized for finger-worn and wrist-worn accelerometers over the range [10^−6^,10^−11^] *g*. The optimal *α* values were determined by maximizing the convergent validity between *r_u_* and the FMA-UE scores. This approach ensured that appropriate thresholds were tailored to accurately define active movements from each sensor location.

### Statistical Analyses of Motor Performance Metrics

We used Spearman correlation coefficients to evaluate the convergent validity of motor performance metrics from finger-worn and wrist-worn sensors with FMA-UE. Moreover, to investigate how different aspects of motor impairments are reflected in the sensor data, we also examined the correlation coefficients with the individual subcomponents of FMA-UE.

To investigate the sensitivity of motor performance metrics in relation to impairment, we divided participants into mildly and moderately impaired groups based on the overall FMAUE score cutoff of 47 (28). In total, we analyzed four subgroups: 1) wrist-derived metrics in the mildly impaired group, 2) finger-derived metrics in the mildly impaired group, 3) wrist-derived metrics in the moderately impaired group, and 4) finger-derived metrics in the moderately impaired group. For within-group comparisons (i.e., comparing finger- and wrist-derived metrics within the same impairment group), the Wilcoxon signed-rank test was used to evaluate the differences between paired samples (29). For between-group comparisons (i.e., comparing metrics derived from the same sensor between the mildly and moderately impaired group), the Mann-Whitney U test was used to evaluate differences between independent samples (30). Effect sizes for between-group comparisons were quantified using Cohen’s *d* (31).

## Results

Twenty-four stroke survivors participated in the study (61.9 ± 9.4 years old; mean ± standard deviation). Table 2 summarizes their demographic and clinical characteristics. Of the 24 participants, 15 were classified as mildly impaired, and 9 as moderately impaired. Four participants used adaptive equipment, such as walkers, to assist with mobility as needed. Data collection averaged 50.0 ± 13.6 minutes per participant.

**Table 2.** The demographic and clinical characteristics of the study participants.

| ID | Age Range (yrs) | Sex | Dominant Side | Affected Side | FMA-UE |
| --- | --- | --- | --- | --- | --- |
| 1 | [66, 70] | Male | Right | Left | 43 |
| 2 | [61, 65] | Male | Right | Left | 55 |
| 3 | [61, 65] | Female | Left | Left | 57 |
| 4 | [76, 80] | Male | Right | Left | 36 |
| 5 | [66, 70] | Male | Right | Right | 44 |
| 6 | [71, 74] | Female | Right | Left | 55 |
| 7 | [66, 70] | Male | Right | Left | 64 |
| 8 | [36, 40] | Male | Right | Right | 53 |
| 9 | [51, 55] | Male | Right | Right | 65 |
| 10 | [56, 60] | Male | Right | Left | 37 |
| 11 | [61, 65] | Female | Right | Right | 61 |
| 12 | [71, 74] | Male | Right | Left | 44 |
| 13 | [41,45] | Male | Right | Right | 59 |
| 14 | [46, 50] | Female | Right | Right | 64 |
| 15 | [61, 65] | Male | Left | Right | 65 |
| 16 | [51, 55] | Female | Left | Left | 60 |
| 17 | [71, 74] | Female | Right | Right | 41 |
| 18 | [66, 70] | Male | Left | Left | 45 |
| 19 | [71, 74] | Male | Left | Left | 46 |
| 20 | [56, 60] | Female | Left | Left | 56 |
| 21 | [66, 70] | Male | Right | Left | 44 |
| 22 | [51, 55] | Male | Right | Right | 51 |
| 23 | [61, 65] | Female | Right | Right | 57 |
| 24 | [56, 60] | Male | Left | Left | 57 |

### Optimizing α for Use Ratio

Figure 2 depicts the variation in Spearman correlation coefficients between FMA-UE scores and use ratios as a function of the acceleration threshold α. The optimal threshold that maximized the correlation coefficient was 9.24 × 10^−3^ *g* for finger-worn accelerometers and 4.18 × 10^−3^ *g* for wrist-worn accelerometers. The higher threshold for the finger-worn device reflects its ability to capture a wider range of motions, including fine hand and gross arm movements, leading to larger signal magnitudes. It is also noteworthy that the optimal threshold for wrist-worn accelerometers (4.18 × 10^−3^ *g*) closely aligns with the established standard of 3.33 × 10^−3^ *g* (23, 27).

**Figure 2.**
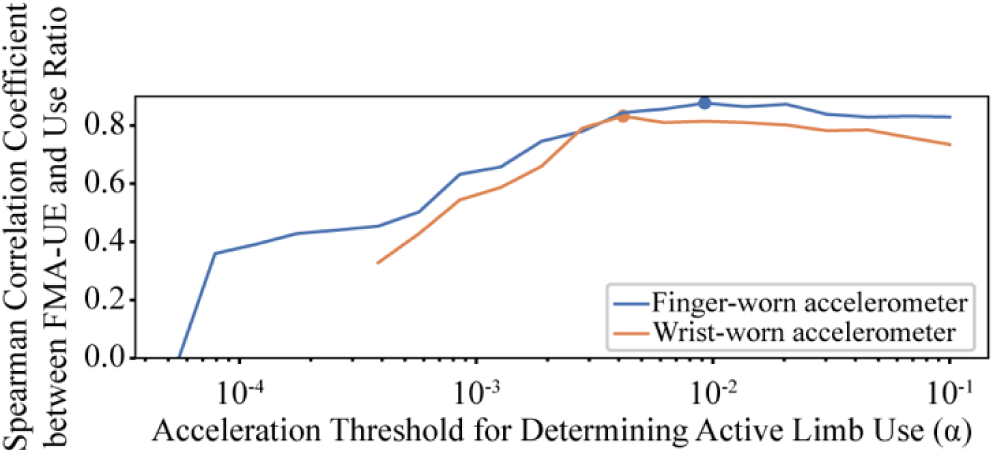
The variation in Spearman correlation coefficients between FMA-UE scores and use ratios derived from finger-worn and wrist-worn accelerometers as a function of the acceleration threshold *α*. The optimal *α* value for each sensor type is marked with a dot.

### Finger-Worn Accelerometers Demonstrate Stronger Convergent Validity than Wrist-Worn Accelerometers

Table 3 summarizes the Spearman correlation coefficients between motor performance metrics and both the overall FMA-UE score and its individual subcomponents. Performance metrics derived from finger-worn accelerometers showed stronger correlations with the overall FMA-UE score compared to those from wrist-worn accelerometers. Moreover, finger-worn sensor metrics exhibited stronger correlations to the wrist and hand components of the FMA-UE, while maintaining comparable correlations to the upper extremity and coordination & speed components. Figure 3 illustrates the relationships between finger sensor-based performance metrics and the overall FMA-UE score; corresponding scatter plots for wrist-based metrics and subcomponent scores are provided in Figures. S1 and S2 in the Supplementary Materials.

**Figure 3.**
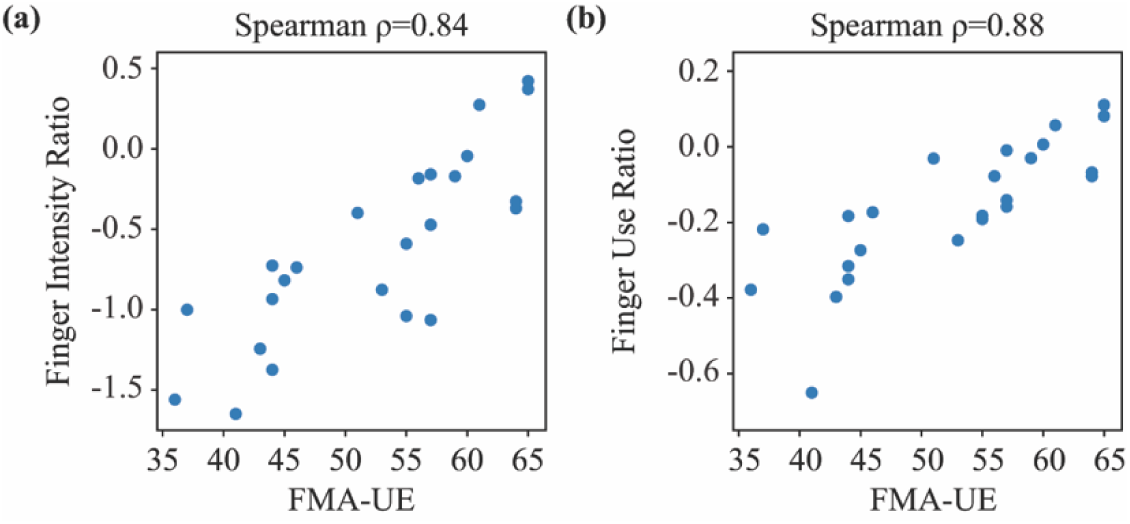
The relationships between the overall FMA-UE score and motor performance metrics from finger-worn accelerometer data: (a) intensity ratio and (b) use ratio.

**Table 3.**
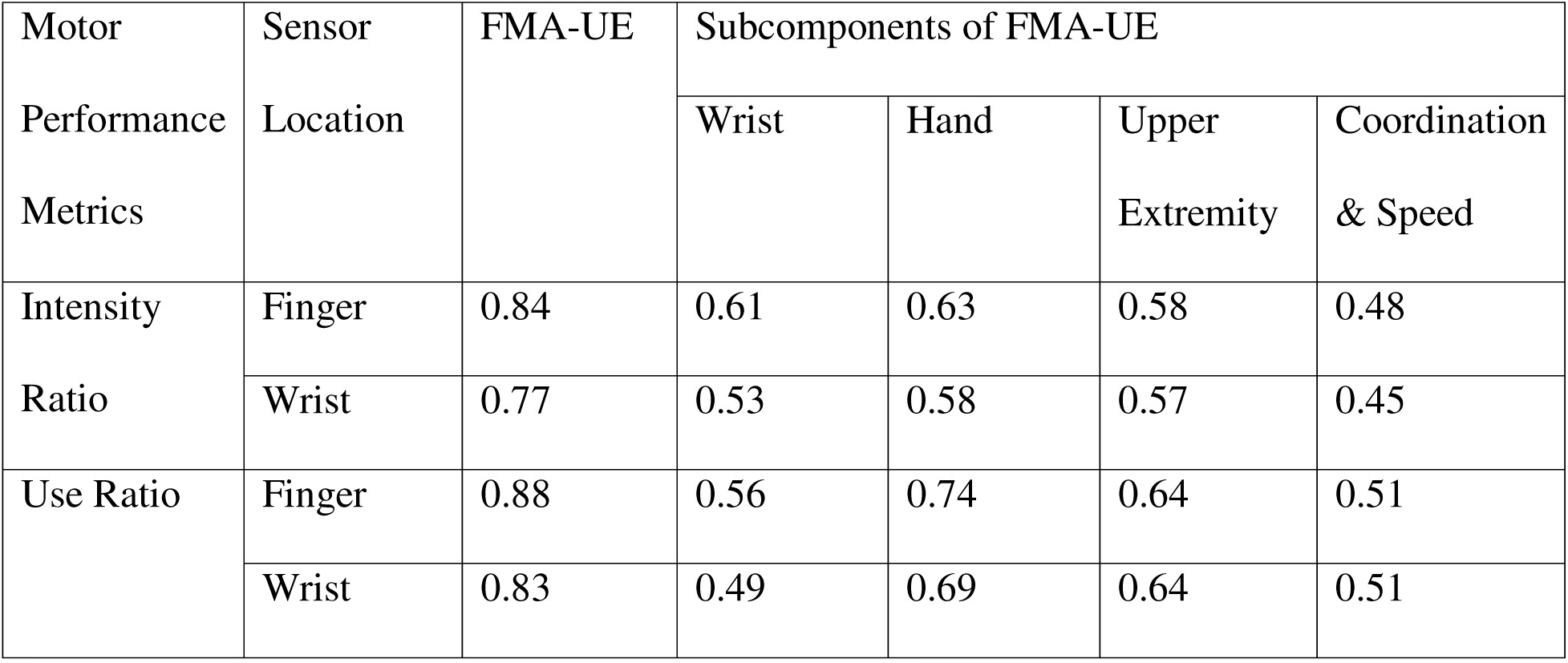
Spearman correlation coefficients between motor performance metrics and overall FMA-UE score and each subcomponent of FMA-UE score.

### Finger-Worn Accelerometers Support Greater Sensitivity in Detecting Subtle Variations than Wrist-Worn Accelerometers

Figure 4 compares metrics from finger-worn and wrist-worn accelerometers between different impairment groups. For the moderately impaired group, the Wilcoxon signed-rank test showed that both mean intensity ratio (*p* < 0.05) and mean use ratio (*p* < 0.01) from finger-worn accelerometers were significantly lower than those from wrist-worn accelerometers, indicating greater sensitivity of finger-worn sensors in detecting asymmetric limb use. In contrast, no significant differences (*p* > 0.05) were observed between sensor locations in the mildly impaired group, although finger-based metrics were slightly lower than those from wrist-worn sensors. Both the intensity ratio and use ratio for this group were closer to zero, reflecting symmetrical movement between the stroke-affected and contralateral limbs.

**Figure 4.**
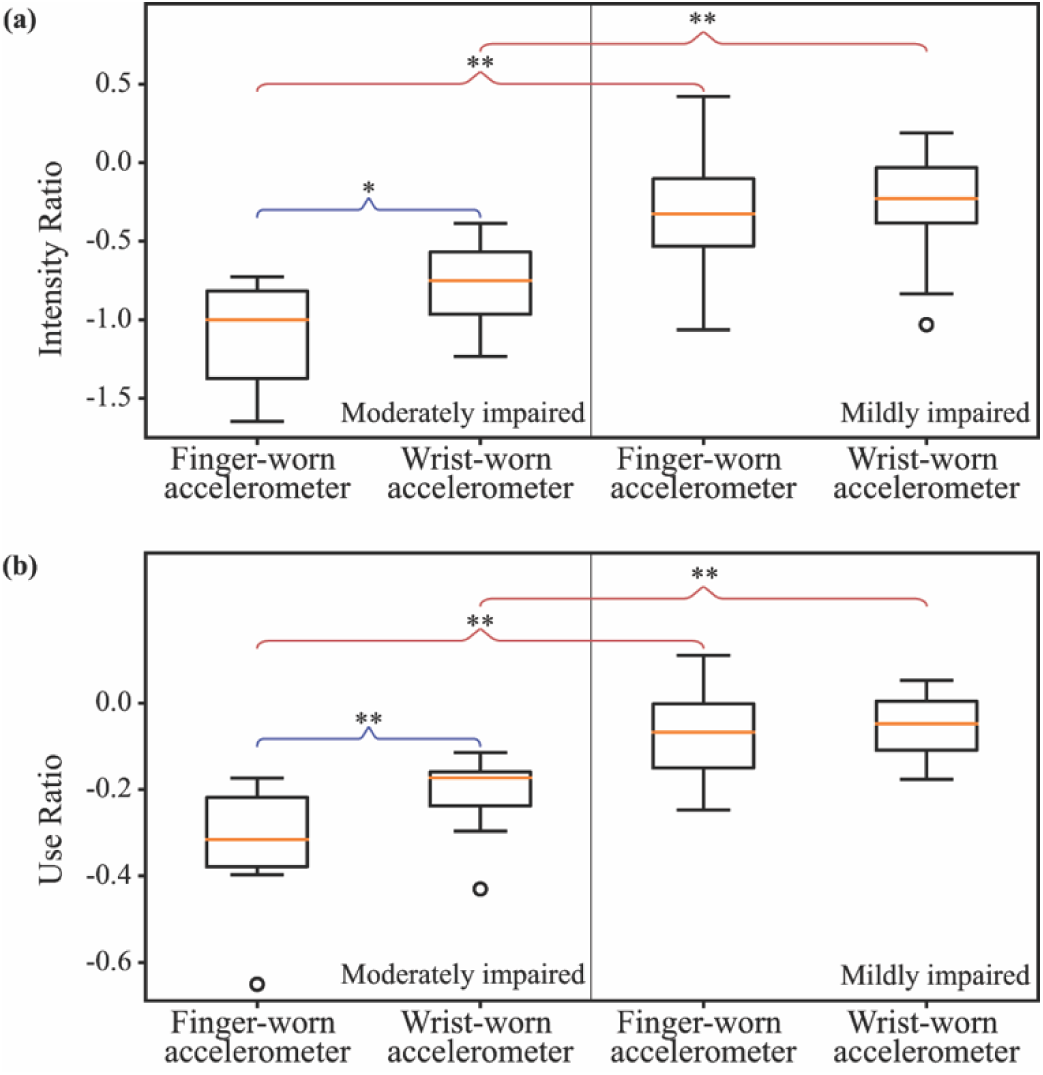
The data were categorized into four groups based on sensor location (finger-worn vs. wrist-worn) and impairment level (mild vs. moderate). Statistical comparisons were performed to identify significant differences between groups. Blue brackets denote significance from the Wilcoxon signed-rank test, and red brackets indicate significance from the Mann-Whitney U test. A single asterisk (*) represents statistical significance at p < 0.05, while a double asterisk (**) indicates p < 0.01.

We also compared motor performance metrics (intensity ratio and use ratio) between mildly and moderately impaired participants for both finger-worn and wrist-worn sensors. Table 4 presents the effect sizes calculated using Cohen’s *d*, revealing that finger-worn sensors consistently yielded larger effect sizes for both metrics compared to wrist-worn sensors. This suggests that finger-worn sensors are more sensitive in detecting motor performance differences related to impairment severity.

**Table 4.**
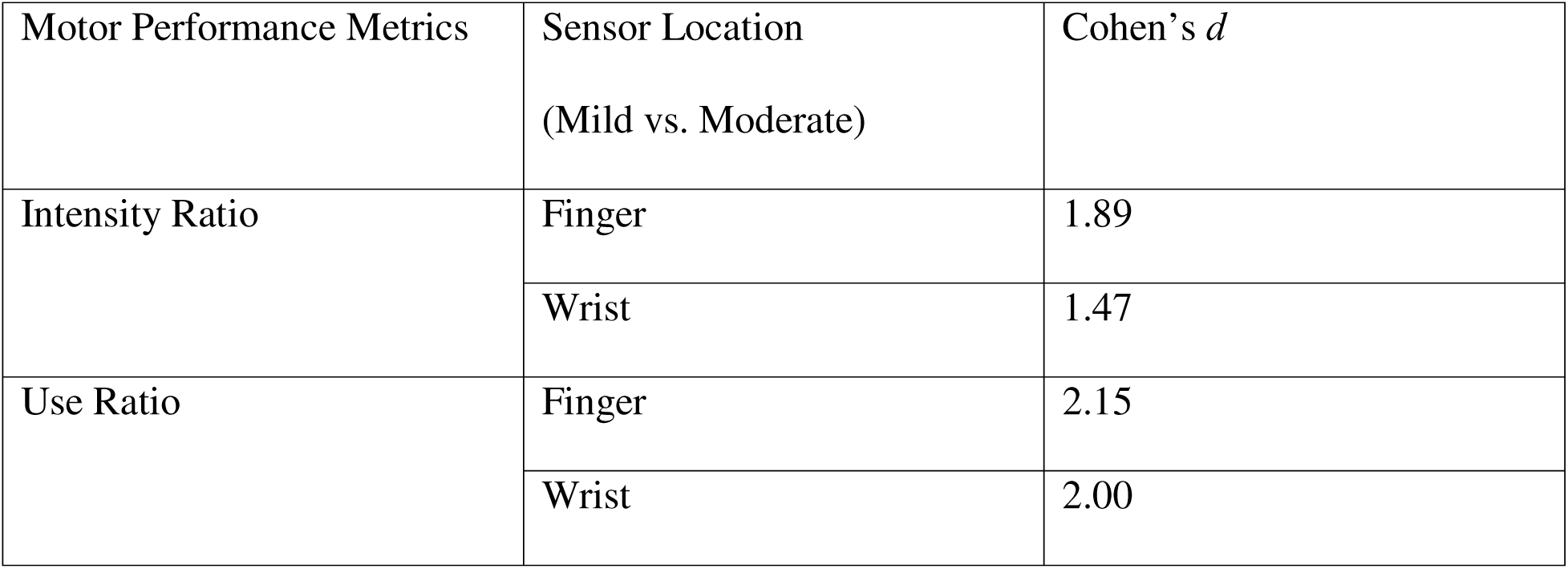
Effect sizes quantifying differences in motor performance metrics between mildly and moderately impaired participants, across the two sensor locations.

## Discussion

This study investigated the efficacy of finger-worn accelerometers for assessing upper-limb motor performance in stroke survivors by analyzing their convergent validity against FMA-UE and its subcomponents. Motor performance metrics derived from finger-worn sensors showed stronger or comparable convergent validity with the overall FMA-UE score and its subcomponents compared to wrist-worn sensors. Additionally, finger-worn accelerometers exhibited greater sensitivity in detecting variations in motor performance across different impairment levels. These findings underscore the potential of finger-worn accelerometers to provide more accurate and clinically relevant assessments of motor performance.

The convergent validity results presented in Table 3 indicate that finger-worn accelerometers offer additional insights into patients’ distal movements, providing a more accurate reflection of their real-world motor performance. For both intensity ratio and use ratio, finger-worn sensors exhibited stronger correlations with the wrist and hand components of the FMA-UE compared to wrist-worn sensors. In contrast, correlations with the upper extremity and coordination & speed components, which primarily reflected proximal arm function, were similar between finger-worn and wrist-worn sensors. This suggests that the enhanced correlations with the overall FMA-UE score for finger-worn sensors stem from their ability to capture distal function. Notably, the correlation coefficients reported here for the overall FMA-UE score (e.g., 0.83 for the use ratio from wrist-worn sensors) exceeded those typically reported in the literature for wrist-worn accelerometers in naturalistic settings, which ranged from 0.59 to 0.67 (32–34). This difference likely arose because our experiment focused exclusively on functional activities requiring upper-limb engagement, whereas real-world scenarios often involved a broader range of activities not specific to upper-limb use (e.g., sleeping and walking) (35). Consequently, our dataset likely contained more targeted information relevant to upper-limb impairment as assessed by the FMA-UE.

As shown in Figure 4, finger-worn sensors demonstrated greater sensitivity in detecting asymmetric limb use, particularly among moderately impaired stroke survivors, revealing an increased reliance on the contralateral hand for both gross and fine motor tasks—a subtle pattern not captured by wrist-worn sensors. Given that the hands and fingers typically recovered later in the rehabilitation process (36), these patients might display more pronounced asymmetries in limb use, which were more effectively detected by finger-worn accelerometers. In contrast, mildly impaired stroke survivors exhibited more symmetric movement intensity and use between the two limbs. This suggested that individuals with mild impairment might have regained functional symmetry in both proximal and distal segments of the affected upper limb, leading to more comparable movement patterns across hands. These findings aligned with previous research demonstrating symmetric upper-limb use in stroke survivors with mild or no impairments, as well as in neurologically intact individuals (14, 37).

We envision finger-worn sensors as valuable tools in outpatient settings to assess patients’ motor performance in their home and community environments. By capturing both gross arm and fine hand movements, these sensors offer comprehensive insights into the use of the stroke-affected limb compared to the contralateral limb, particularly for individuals with moderate upper-limb impairments. Such data could support clinicians in designing targeted therapies to enhance patients’ motor performance (8, 9). Previous studies indicate that stroke survivors are receptive to wearing ring-form-factor devices for prolonged periods, and clinicians are open to integrating sensor data into routine practice to better understand real-world motor performance and facilitate collaborative therapy goal-setting (15). The growing popularity of commercial ring-based health monitoring devices—such as the Oura Ring and Samsung Galaxy Ring— further supports their potential acceptability. Nevertheless, assessing their long-term acceptability in this population remains a critical area for future investigation.

This study has several limitations. First, the small sample size (N = 24) First, the sample size (N = 24) is smaller than the number recommended by the COSMIN guidelines, which suggest at least 50 participants for validating outcome measurement instruments (38, 39). Future studies should recruit a larger cohort of stroke survivors to increase statistical power and further enhance the generalizability of the findings. Second, although participants performed ADLs in a simulated environment designed to mimic natural movements, the data may not fully reflect their real-world motor performance. To enhance validity, future studies should recruit a larger, more diverse cohort with a wider range of upper-limb motor impairments and collect data in naturalistic settings. Moreover, the acceleration threshold *α* used to calculate use ratios was optimized on the current dataset without validation on an independent dataset or through cross-validation techniques. Future studies should evaluate the robustness of *α* across diverse populations and settings to enhance the generalizability of the findings. Finally, further research is needed to assess whether the proposed upper-limb performance metrics are sensitive to longitudinal changes in motor capacity. Prior studies have indicated that limb activity measures from wrist-worn accelerometers often lack responsiveness to such changes (13, 40), which could be due to their limited ability to detect fine motor improvements. Given the stronger alignment of our proposed metrics with clinical motor capacity assessments, we hypothesize that they may demonstrate greater responsiveness.

## Conclusion

This study comparatively investigated finger-worn accelerometers, which capture both gross arm and fine hand movements, as an assessment tool for post-stroke upper-limb motor performance, compared to wrist-worn accelerometers. The findings demonstrate that finger-worn accelerometers exhibit stronger convergent validity and greater sensitivity in detecting subtle variations in motor performance compared to established wrist-worn devices. These results lay the groundwork for establishing a more precise and objective link between motor capacity and performance in stroke survivors, paving the way for improved rehabilitation assessments and personalized therapeutic interventions.

## Data Availability

All data produced in the present work are contained in the manuscript and not available online

## Acknowledgements

The authors would like to thank the research staff in the Motion Analysis Laboratory at Spaulding Rehabilitation Hospital for their invaluable assistance in conducting data collection among stroke survivors.

## Ethical Approval

The study was approved by the Institutional Review Board of Mass General and Brigham Hospital (IRB #2019P002612).

## Declaration of Conflicting Interests

The author(s) declared no potential conflicts of interest with respect to the research, authorship, and/or publication of this article.

## Funding

The author(s) disclosed receipt of the following financial support for the research, authorship, and/or publication of this article: This work was supported in part by the National Institute of Biomedical Imaging and Bioengineering (grant number R01EB027777).

## Notes

### Competing Interest Statement

The authors have declared no competing interest.

### Author Declarations

Ethics committee/IRB of Mass General and Brigham Hospital gave ethical approval for this work (IRB #2019P002612)

