## Supplemental materials for "Beyond the Wrist: Finger-Worn Accelerometers Enhance Assessment of Post-Stroke Motor Performance"

**Supplementary Materials**

Figure S1 and Figure S2 illustrate the relationships between overall FMA-UE score and each

component of FMA-UE score with sensor-based performance metrics for finger-worn data and

wrist-worn data, respectively.

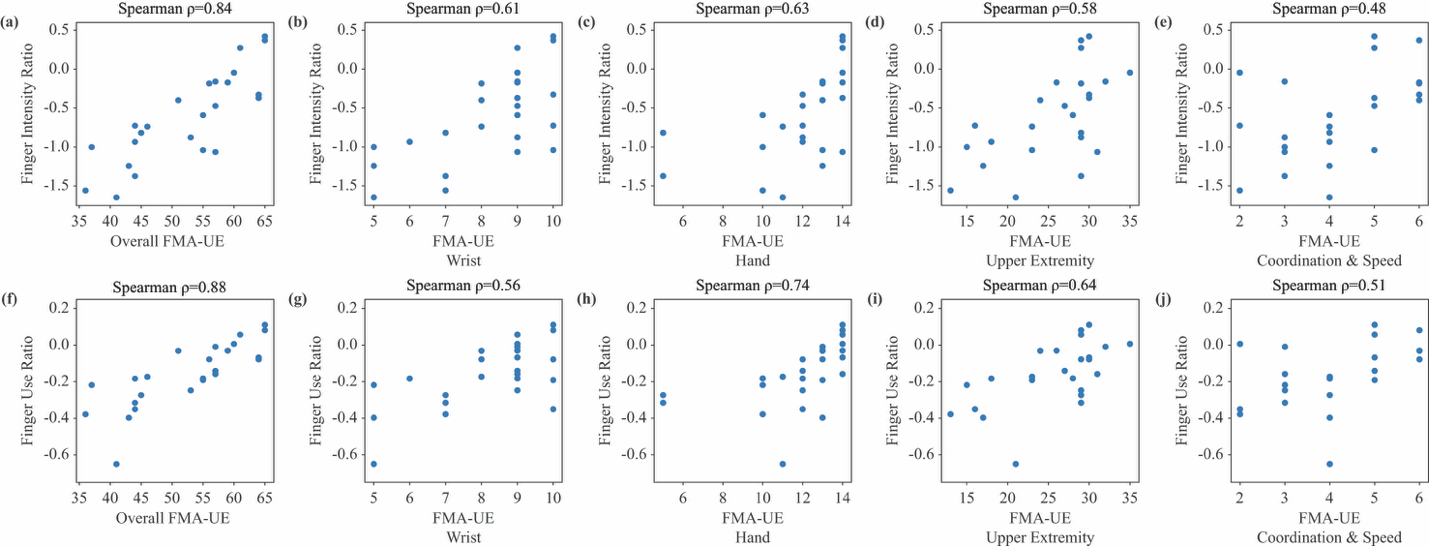

Figure S1. The relationships between the overall FMA-UE score and each component of FMA-UE score with motor performance metrics from finger-worn accelerometer data: (a)-(e) intensity ratio and (f)-(j) use ratio.

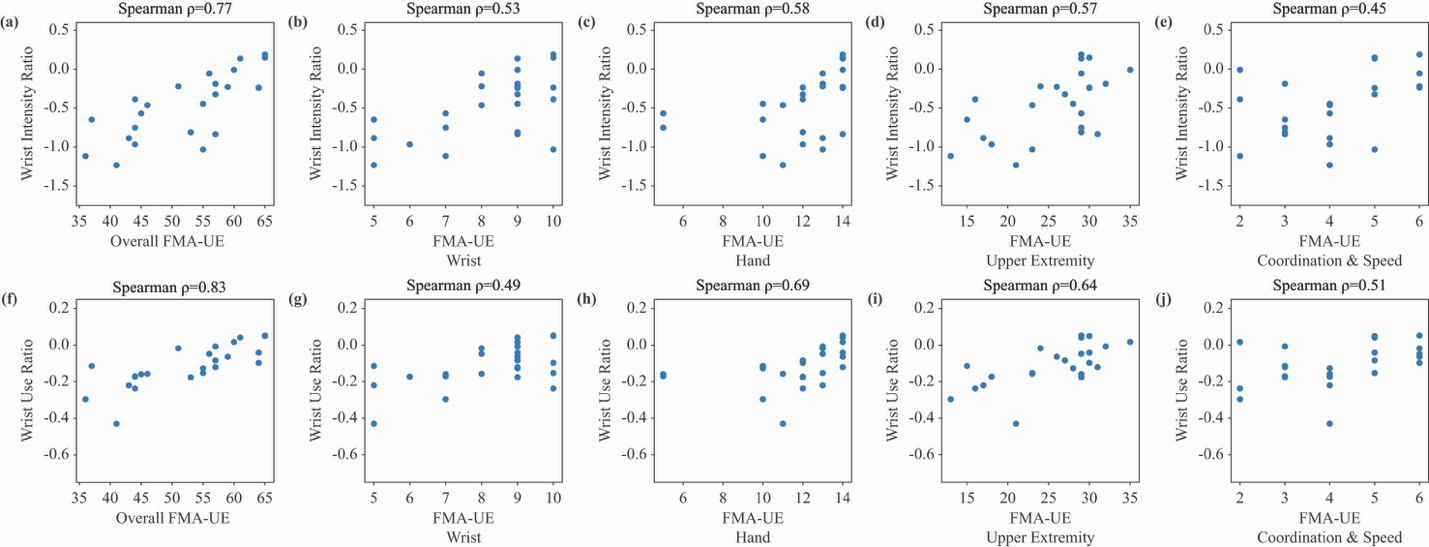

Figure S2. The relationships between the overall FMA-UE score and each component of FMA-UE score with motor performance metrics from wrist-worn accelerometer data: (a)-(e) intensity ratio and (f)-(j) use ratio.
